# Fluid and White Matter Suppression Contrasts MRI Improves Deep Learning Detection of Multiple Sclerosis Cortical Lesions

**DOI:** 10.1101/2025.01.19.25320792

**Authors:** Pedro M. Gordaliza, Jannis Müller, Alessandro Cagol, Nataliia Molchanova, Francesco La Rosa, Charidimos Tsagkas, Cristina Granziera, Meritxell Bach Cuadra

## Abstract

**Purpose:** To investigate, for the first time, the efficacy of Fluid and White Matter Suppression (FLAWS) MRI sequence in improving Deep Learning (DL)-based detection and segmentation of cortical lesions in Multiple Sclerosis (MS) patients even, with applicability to clinical settings where only standard T1-weighted images are available.

**Materials and Methods:** In this retrospective multi-site study, we analyzed 204 MS patients using DL models developed with FLAWS and Magnetization Prepared 2 Rapid Acquisition Gradient Echoes (MP2RAGE) sequences. Reference standard annotations were established through two approaches: (1) consensus of three expert raters across all contrasts, and (2) single-rater annotations for individual modalities. Models were validated on both internal and external datasets, with performance assessed using *F*_1_-score for detection and DSC for segmentation accuracy.

**Results:** Models involving FLAWS demonstrated superior performance over MP2RAGE-only models. The combined MP2RAGE+FLAWS model achieved CL detection with median *F*_1_-score of 0.667[0.339*−*0.840] compared to multirater consensus. Models trained on comprehensive consensus annotations outperformed those trained on single-modality annotations. Notably, a model exclusively based on MP2RAGE images and trained with FLAWS-derived annotations showed, showed strong generalization to external Magnetization Prepared Rapid Gradient-Echo (MPRAGE) clinical datasets (median *F*_1_-score: 0.55[0.211 *−* 0.998]).

**Conclusion:** Integration of FLAWS-derived contrasts and annotations significantly improves DL-based CL detection and segmentation. The models demonstrate capability in identifying lesions missed by individual raters and maintain robust performance even without FLAWS sequences in standard clinical settings. This advancement facilitates clinical translation, supported by publicly available inference models on DockerHub.

**Graphical Abstract:** 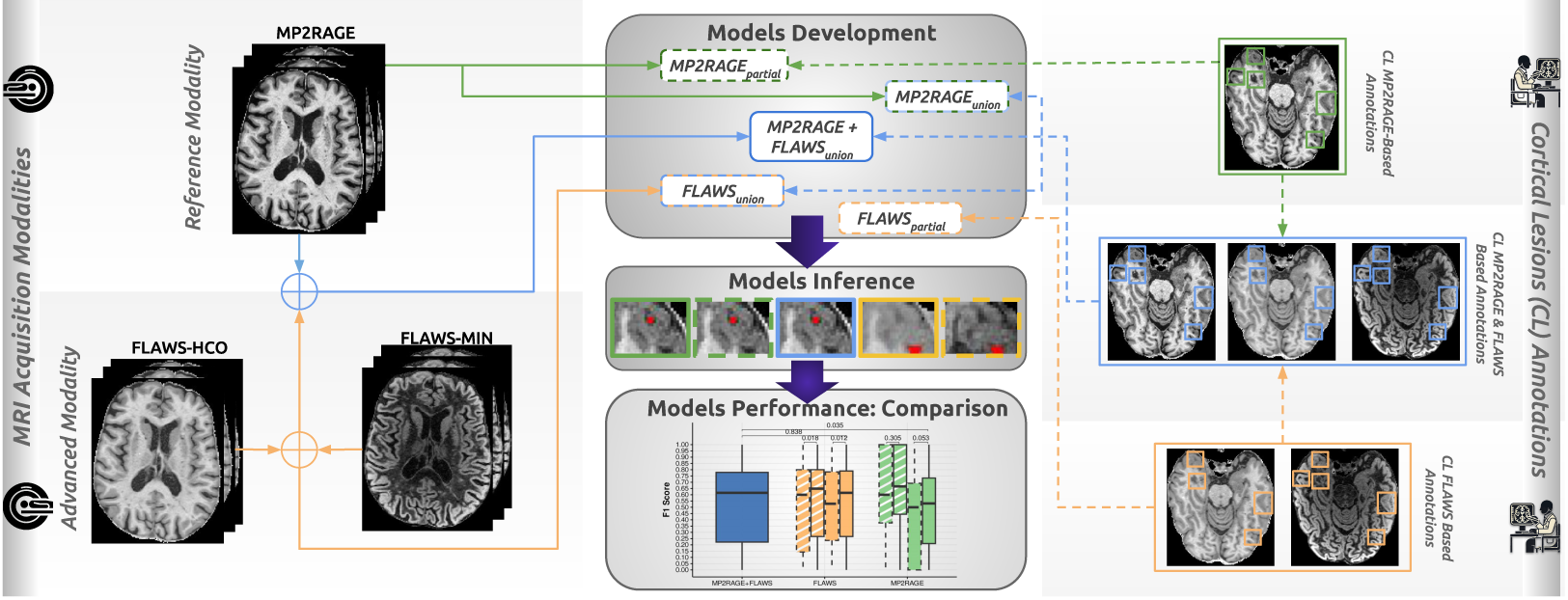

**Highlights:**

- MP2RAGE+FLAWS MRI sequences achieves superior cortical lesion detection performance
- Quantitative validation shows median *F*_1_ of 0.667[0.339 *−* 0.840] with combined sequences
- FLAWS-trained model generalizes well to standard MPRAGE images (*F*_1_: 0.55[0.211 *−* 0.998)
- Model demonstrates immediate clinical applicability on standard MPRAGE without FLAWS input
- Implementation publicly accessible via DockerHub for widespread clinical adoption

## 1. Introduction

Magnetic Resonance Imaging (MRI) is pivotal for profiling neurological disorders and indispensable for assessing those with complex etiology, such as Multiple Sclerosis (MS) [1, 2]. Disease-related imaging biomarker assessment provides insights into the underlying biological mechanisms driving these conditions. In MS, the most commonly employed imaging biomarkers are White Matter Lesions (WML), visualized as hyperintensities on T2-weighted contrasts. Despite their lack of sensitivity, WML dissemination in space and time is required for MS diagnosis according to *McDonald criteria* [1, 3]. Recent expert consensus, however, has indicated Cortical Lesions (CL) can provide increased specificity, as evidenced using advanced MRI techniques [4, 5, 6], verified through histopathology studies [7]. However, the inclusion of CL in routine assessments remains limited, primarily due to a lack of sensitivity in detecting these using conventional MRI scanners and sequences. Enhancing CL detection necessitates the utilization of dedicated advanced multi-contrast MRI sequences and/or high-field scanners, offering greater sensitivity in gray matter and a finer spatial resolution [8]. However, data of both natures are limited, hindering the development and application of Deep Learning (DL) methods [9] to assist in the more clinically relevant identification of CL. The cost and time constraints associated with ultra high-field scanners confine their use to few research settings, where experts manually identify CL for specific trials, albeit with moderate inter-rater variability (i.e., Cohen *κ ≈* 0.50 *−* 0.69) [7].

Leveraging new advanced MRI sequences using clinical scanners offers a more widely accessible solution, despite shared challenges such as limited sensitivity, time-consuming manual evaluation and data scarcity. Contrasts such as Magnetization Prepared 2 Rapid Acquisition Gradient Echoes (MP2RAGE) (*κ* = 0.582 in [10]), Phase Sensitive Inversion Recovery, Magnetization Prepared Rapid Gradient-Echo (MPRAGE) or Double Inversion Recovery (DIR) (*κ* = *−*0.36 in [11]) have been assessed to this end.

A promising new sequence in this context is Fluid and White Matter Suppression (FLAWS) [12]. FLAWS provides both white matter and cerebrospinal fluid-suppressed 3D high spatial resolution contrasts simultaneously in one acquisition (*FLAWS1* and *FLAWS2*). These signals can be combined in a voxel-based manner to reconstruct multiple contrasts (i.e., FLAWS*_HCO_* for the high contrast images after WM signal suppression and FLAWS*_MIN_* for the minimum of both FLAWS. See Figure 1 and Figure 5), showing higher sensitivity than MP2RAGE, e.g., in [10] its reported that FLAWS*_HCO_* and FLAWS*_MIN_* detect in median per patient 4.5 CL compared to 3 detected on MP2RAGE (p-value *<* 0.001).

**Figure 1:**
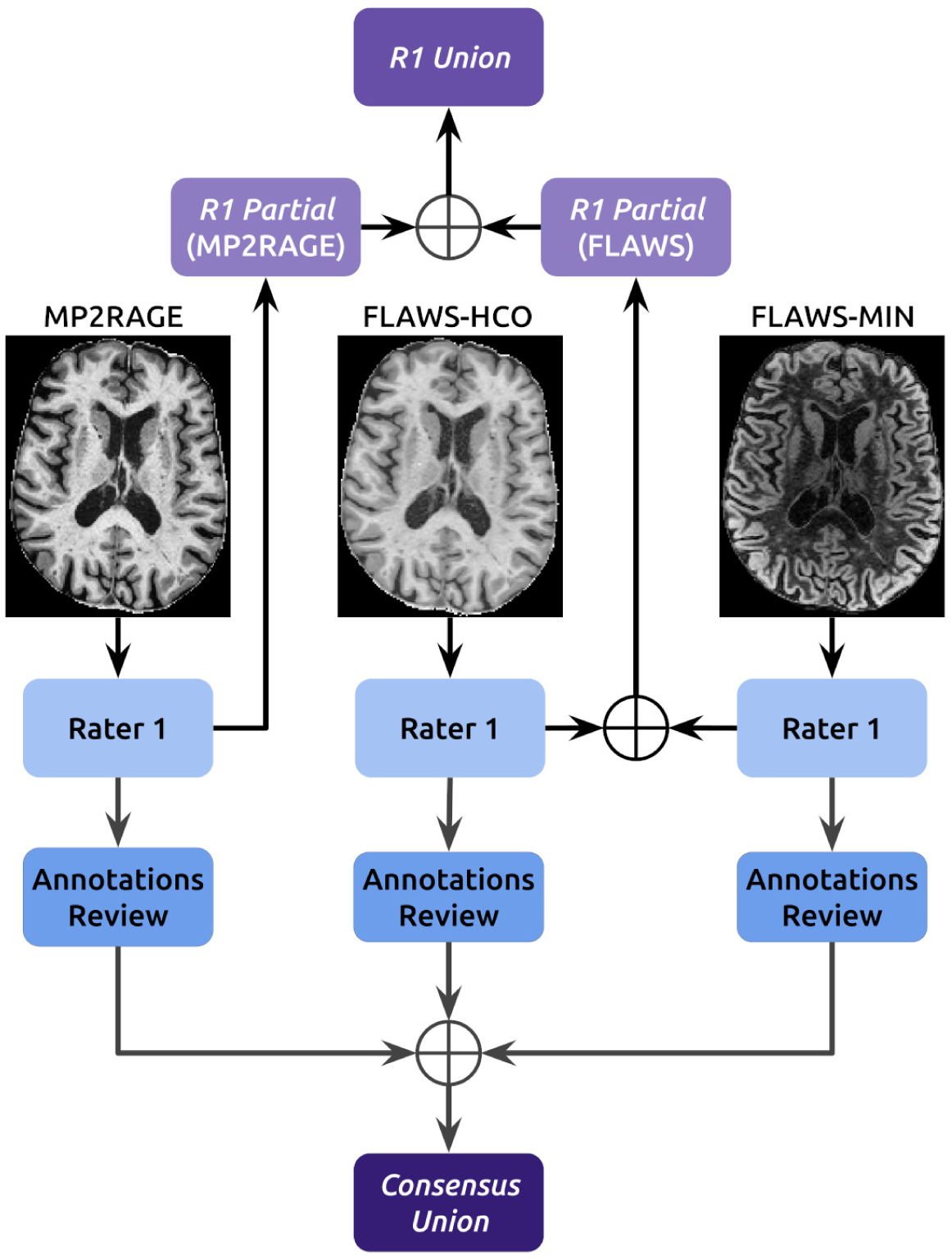
Schematic representation of the reference standard annotation process for *MP2RAGE and FLAWS* dataset. Three MRI sequences (Magnetization Prepared 2 Rapid Acquisition Gradient Echoes, Fluid and White Matter Suppression-HCO, and FLAWS-MIN) are independently annotated by Rater 1 (*R1*). These annotations are combined to form *R1 partial* annotations for MP2RAGE and FLAWS separately, and *R1 Union* annotations combining all sequences. The initial annotations of 30 subjects undergo review, leading to the *Consensus Union* employed for Experiment 2

Artificial Intelligence (AI) methods have shown promise in easing the task of lesion detection. However, these efforts have predominantly focused on WML, with only a few attempts to address CL detection, either along with WML under the general lesion category [13] or using 7T MRI [14]. This partly explains the modest integration of automated tools into MS clinical pipelines [15].

These promising findings steer the aim for our work: for the very first time, assess the capabilities of state-of-the-art DL models when employing the FLAWS sequence for CL detection and segmentation. Our approach focuses on the following objectives: 1) To evaluate how the inclusion of FLAWS sequence enhances CL automatic assessment compared to using MP2RAGE alone, 2) To assess the model perfomance in correctly identifying false negative and false positives findings in single-rater annotations, and 3) To investigate the capability of DL models trained on FLAWS-derived information to assess CL when only common research sequences (e.g., MP2RAGE) or standard clinical sequences (e.g., MPRAGE) are available, measuring the model’s ability to perform accurately when the test data differs (shift) from the training data.

By addressing these objectives, we aim to rigorously evaluate the performance of DL models in a low data regime as often presented in clinical contexts. Our aim is to support the early integration of these advanced DL models in clinical research settings, enhancing CL assessment by medical doctors, and, consequently, improving MS diagnosis and monitoring.

## 2. Materials and Methods

For this study, institutional review board approval and written informed consent were obtained from all participants prior to enrollment in the study.

### 2.1. Study design

This retrospective, exploratory study evaluates the feasibility of leveraging FLAWS-derived contrasts for automatic CL detection and segmentation in MS, aiming to enhance diagnosis and disease monitoring.

This study uses the state-of-the-art *nnU-Net* framework v2.0 [16] for semantic segmentation, subsequently instantiated to obtain individual CL. We develop DL models for various contrasts and their derived annotations combinations. Through comparative analysis, we assess the impact of FLAWS-derived contrasts on automated CL assessment in MS patients, evaluating their effectiveness both when these contrasts are directly accessible and when applying FLAWS-trained models to non-FLAWS data.

### 2.2. Dataset Description

MRI scans from 204 MS patients were included. The scans were acquired at two centers: University Hospital Basel (hereafter Hospital A; N=163) between 2020-2022 and Lausanne University Hospital (hereafter Hospital B; N=41) between 2017-2019. Imaging was performed on *Prisma* (Hospital A) and *Trio* (Hospital B) 3.0 T MRI scanners (Siemens Healthineers, Erlangen, Germany). All patients met the 2017 McDonald criteria [3] for MS diagnosis. Detailed cohort characteristics and imaging protocols are summarized in Table 1.

**Table 1:** Summary of the available datasets. Values expressed as mean ± standard deviation or median [interquartile range]. EDSS: Expanded Disability Status Scale. RRMS: Remitting Relapsing Multiple Sclerosis. SPMS: Secondary Progressive Multiple Sclerosis. PPMS: Primary Progressive Multiple Sclerosis

| Dataset | Hospital A (Basel) |  | Hospital B (Lausanne) |  |
| --- | --- | --- | --- | --- |
|  | *A-MP2RAGE and FLAWS* | *A-MP2RAGE* | *B-MP2RAGE* | *B-MPRAGE* |
| **Data Partition (N)** | Train.(39), Internal test.(30) | Internal test. (94) | External testing (41) |  |
| **Female** | 41 (51.9%) | 56 (59.6%) | 25 (61%) |  |
| **Age** | 43.4 ± 13.6 | 45.76 ± 14.15 | 34 ± 9 | |
| **Disease type: RRMS/SPMS/PPMS** | 41/19/9 | 56/25/13 | 26/8/7 |  |
| **EDSS** | 2.5 [1.5-4] | 3 [2-4.55] | 1.5 [1.5-2] |  |
| **Dis. duration [y]** | 6.40 [1.23-16.03] | 6.40 [0.91-17.05] | 27 [7.50-41.25] |  |
| **CLs per patient** | 9 [1-19] | 2 [0-6.75] | 3 [1-9] |  |
| **MRI Scanner** | Siemens Prisma (3T) |  | Siemens Trio (3T) |  |
| **Acquisition Dates** | 2018 - 2022 |  | 2017 - 2019 |  |
| **Sequences** | FLAWS | MP2RAGE | MP2RAGE | MPRAGE |
| **Resolution [$mm^3$]** | 1×1×1 | | 1×1×1.2 | 1×1×1.2 |
| **Orientation** | Sagittal |  | Sagittal |  |
| **FOV** | 256×240×192 | 256×240×176 | 256×240×176 | 256×240×160 |
| **TR/TE/T1/T2 (ms)** | 5000/2.19/449/1270 | 5000/2.98/700/2500 | 5000/2.89/700/2500 | 2300/2.98/900/- |

The dataset was divided into four subsets:

- Internal dataset
  1. *A-MP2RAGE and FLAWS* (N=69, Hospital A): Includes both FLAWS sequence (with 3D-derived image contrasts: FLAWS*_HCO_* and FLAWS*_MIN_*) and MP2RAGE. This subset serves as internal data for training and validation (N = 39) and testing (N = 30). Stratification was carried out to keep the same CL prevalence and balance sex and age.
  2. *A-MP2RAGE* (N=94, Hospital A): Comprises patients with only MP2RAGE scans, used for internal testing.
- External test dataset from conventional research and clinical sequences (N=41, Hospital B).
  3. *B-MP2RAGE* : This external testing dataset comprises scans acquired using an early implementation of the MP2RAGE sequence. The acquisition parameters of this protocol differ from the standardized parameters commonly employed in current practice.
  4. *B-MPRAGE* : Clinically employed MPRAGE scans.

### 2.3. Image Preprocessing

In the *MP2RAGE and FLAWS* dataset, MP2RAGE images for each patient were linearly registered to the corresponding FLAWS*_HCO_* using *Advanced Normalization Tools* (ANTs) v.2.3.1 [17]. All datasets underwent skull stripping using SynthStrip v2.0 [18]. In addition, within the *nnU-Net* framework z-score normalization was automatically performed.

### 2.4. Reference Standard: Cortical Lesions Annotations

All images were manually annotated by expert neurologists or neuroradiologists using ITK-SNAP [19]. The raters were blinded to the clinical status of the patients, and the CL delimitation followed the guidelines from [20, 10]. CL in the *MP2RAGE and FLAWS* dataset were manually segmented by J.M., 6 years of experience, (*R1*) on MP2RAGE, FLAWS*_HCO_*, and FLAWS*_MIN_*. This approach enabled the creation of two types of reference standards for subsequent model development and evaluation (see Figure 1 upper path): *R1 union*, corresponding to the union of segmentations from all three sequences, and *R1 partial*, derived from a subset of sequences—either solely from MP2RAGE annotations or from the combined FLAWS*_HCO_* and FLAWS*_MIN_* annotations always obtained independently on each contrast. To assess model capabilities to automatically detect overlook and misidentified CL, the segmentations of 30 out of the 69 patients in the dataset were reviewed by two additional experts (C.T. and A.C., with 6 and 8 years of experience, respectively), resulting in a final consensus segmentation map for each modality subsequently merged (*Consensus union*).

For the *A-MP2RAGE* dataset, manual segmentation was performed by A.C. based exclusively on the MP2RAGE contrast.

For external testing data in Hospital B (i.e., *B-MP2RAGE* and *B-MPRAGE*), C.G. 22 years of experience conducted the manual segmentation using MP2RAGE.

### 2.5. Model Development

We developed, trained and evaluated five segmentation-supervised models using data from 39 subjects (19.12% of total cohort) with both MP2RAGE and FLAWS sequences available. Our model development strategy explored the effectiveness of different sequence combinations and annotation approaches, reflecting common scenarios where novel sequences are initially tested on limited, partially annotated datasets.

The models varied in their input sequences and annotation sources:

1. *MP* 2*RAGE* + *FLAWS_union_*: Both FLAWS contrasts and MP2RAGE, with *R1 union* annotations
2. *FLAWS_union_*: Only FLAWS contrasts, with *R1 union* annotations
3. *FLAWS_partial_*: Only FLAWS contrasts, with *R1 partial (FLAWS)* annotations
4. *MP* 2*RAGE_union_*: Only MP2RAGE, with *R1 union* annotations
5. *MP* 2*RAGE_partial_*: Only MP2RAGE, with *R1 partial (MP2RAGE)* annotations

Table 2 summarizes each model’s configuration and its application in the experiments detailed in Experimental Design, which align with our study objectives.

**Table 2:**
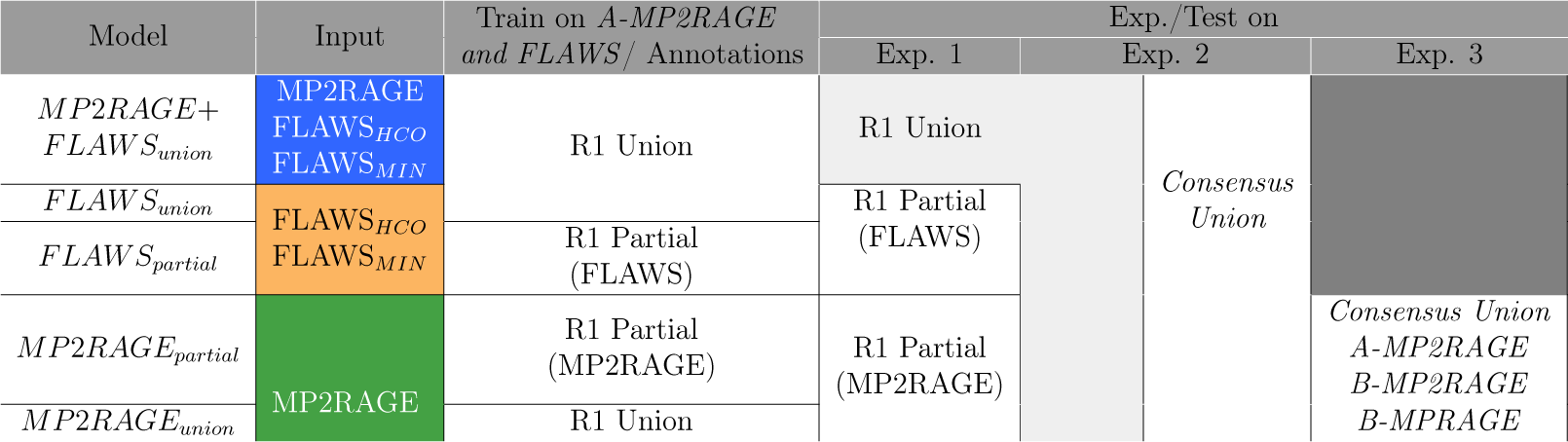
Summary of model configurations, training data, and their application across experiments. Each experiment corresponds to a specific study objective: Experiment 1 evaluates FLAWS contribution to CL assessment (Objective 1), Experiment 2 assesses the detection of single-rater false positive and negative findings (Objective 2), and Experiment 3 investigates CL assessment using common clinical sequences employing models trained with FLAWS annotations (Objective 3).

Each model was built using the *nnU-Net* framework [16], which automatically configures image intensity normalization during preprocessing (see Image Preprocessing), network architecture and training. No additional postprocessing was required for any model. For all models, the *3D full-resolution* architecture was employed.

The training process involved:

- Extensive data augmentation (details in [16] Supplementary Materials 4).
- 1000 training epochs for five-folds, each with different model weight initializations to enhance generalization.
- Post-training ensemble creation by voxel-wise averaging of softmax probabilities from the final layer across the five models.
- Final binary semantic segmentation obtained by thresholding the ensemble’s voxel-wise softmax probabilities, classifying voxels based on the channel (Cortical Lesions or rest) with the highest probability.

To further refine the training process, we modified the *nnU-Net* learning strategy by:

- Employing the ADAM optimizer with an initial learning rate of 3 *×* 10*^−^*^4^ [21].
- Implementing *Blob Loss* [22], which is more appropriate for the sparse nature of CL, in place of the default loss function in *nnU-Net*.
- Utilizing a deep supervision strategy with *Blob Loss*, combining Dice Similarity Coefficient (DSC) loss [23] and Cross Entropy loss for each detected blob.

All models were trained using an NVIDIA RTX3090 (24Gb vRAM) GPU. To ensure reproducibility and facilitate utilization, the models are available on DockerHub.

### 2.6. Experimental Design

We conducted three main experiments to address our objectives:

1. We evaluated whether the FLAWS sequence enhances automatic CL detection, both when used as model input (alone or combined with MP2RAGE) and when the model is trained on annotations derived from it (*R1 union* or *R1 partial*). This experiment utilized 30 held-out subjects for internal testing from Hospital A not included in the FLAWS-derived contrasts training set.
2. Evaluation of the models’ ability to improve CL detection accuracy by comparing their performance against expert manual consensus. This experiment assesses the models’ capacity to identify CL overlooked by a single rater and to correctly exclude false positives initially misidentified as CL. The assessment compares the models’ predictions (trained on *R1 union* annotations) against both single-rater (*R1 union*) and expert consensus (*consensus union*) reference standard annotations for the 30 internal testing subjects.
3. To assess the transferability of knowledge, we evaluated the performance of models trained without FLAWS contrasts as inputs but utilizing

FLAWS-derived annotations (i.e., *MP* 2*RAGE_union_* and *MP* 2*RAGE_partial_*models). These models were applied to MP2RAGE, a more widely available contrast than FLAWS, and MPRAGE, one of the clinically recommended contrast for MS CL detection. This experiment utilized the complete *A-MP2RAGE* dataset (N=94 subjects from the internal dataset) and two external testing datasets from Hospital B (N=41 subjects), which exhibited significant distributional shifts compared to the training data (see Table 1) [24]. These external datasets included *B-MP2RAGE* (MP2RAGE sequence acquired on a different scanner with minor protocol variations) and *B-MPRAGE* (MPRAGE sequence with distinct acquisition protocol and scanner parameters).

### 2.7. Evaluation and Statistical Analysis

Model evaluation adhered to the *Metrics Reloaded* framework [25], treating the task as instance segmentation to account for the sparse nature of CL. We instantiated reference standard annotations and predictions into CL blobs using the Connected Component algorithm with 18-connectivity [26]. To match the reference and predict CL, we employed a Mask Intersection over Union (IoU) threshold of 10% and the Greedy by Score assignment strategy. For all experiments, performance assessment focused on two key metrics: the *F*_1_-score for detection and the DSC for overlap between CL positive instances. The *F*_1_ score, which ranges from 0 to 1, provides a balanced measure of the model’s precision (correctness of positive predictions) and recall (ability to detect all positive cases). A higher *F*_1_-score indicates better overall detection performance. These metrics are summarized using boxplots to illustrate the distribution of performance across different models and conditions.

Statistical analysis for all experiments included paired Wilcoxon signedrank tests to assess differences between models in averaged per-patient *F*_1_ and DSC. To control for multiple comparisons, we applied the Benjamini-Hochberg correction.

Additionally, for each experiment, we conducted correlation analyses and created corresponding Bland-Altman plots. For Experiments 1 and 3, we analyzed the correlation between the number of detected lesions and the reference standard. Bland-Altman plots were used to assess agreement and potential biases in lesion count detection. For Experiment 2, we performed correlation analysis on the *F*_1_ scores obtained using single-expert annotations versus consensus annotations. The corresponding Bland-Altman plot evaluated the differences in *F*_1_ scores between these two annotation methods, providing insight into the consistency and potential biases in model performance across different reference standards. The choice between Pearson or Spearman correlation coefficients was based on the normality of data distribution in each case. All statistical analyses and visualizations were conducted using R (version 4.1.2) [27] and *rstatix* (version 0.7.2) [28].

## 3. Results

Table 1 summarizes demographic characteristics for all datasets. Results are shown as mean *±* standard deviation or median [interquartile range].

### 3.1. Experiment 1

This experiment evaluated the performance of models trained with either FLAWS, MP2RAGE or jointly FLAWS and MP2RAGE sequences as inputs, using both partial and union annotations. Results are presented in Figure 2. Analysis of *F*_1_ scores (Figure 2 top-left) revealed that models trained with annotations from all contrasts (*R1 union*, solid contour lines) consistently outperformed those trained with partial annotations (dashed contour lines), regardless of input sequences and reference annotations. Among models trained with *R1 union* annotations, using only FLAWS sequences, *FLAWS_union_* (solid orange boxplot with solid linetype), demonstrated superior detection performance compared to the model using only MP2RAGE, *MP* 2*RAGE_union_*, (median *F*_1_ = 0.611 *±* 0.36 vs. *F*_1_ = 0.531 *±* 0.36, respectively). The highest detection performance was achieved by the combined *MP* 2*RAGE* + *FLAWS_union_* model (median *F*_1_ = 0.615 *±* 0.26), showing a statistically significant improvement over the *MP* 2*RAGE_union_* model (*p* = 0.036). Note that the results tested against partial annotations (represented by stripped patterns in Figure 3) are only comparable within the same sequence type (i.e., FLAWS or MP2RAGE). These partial annotation results should not be compared across different sequences or with the union annotation results, as they represent different subsets of the total lesion load. The union annotation results (solid patterns), however, are comparable across all models.

**Figure 2:**
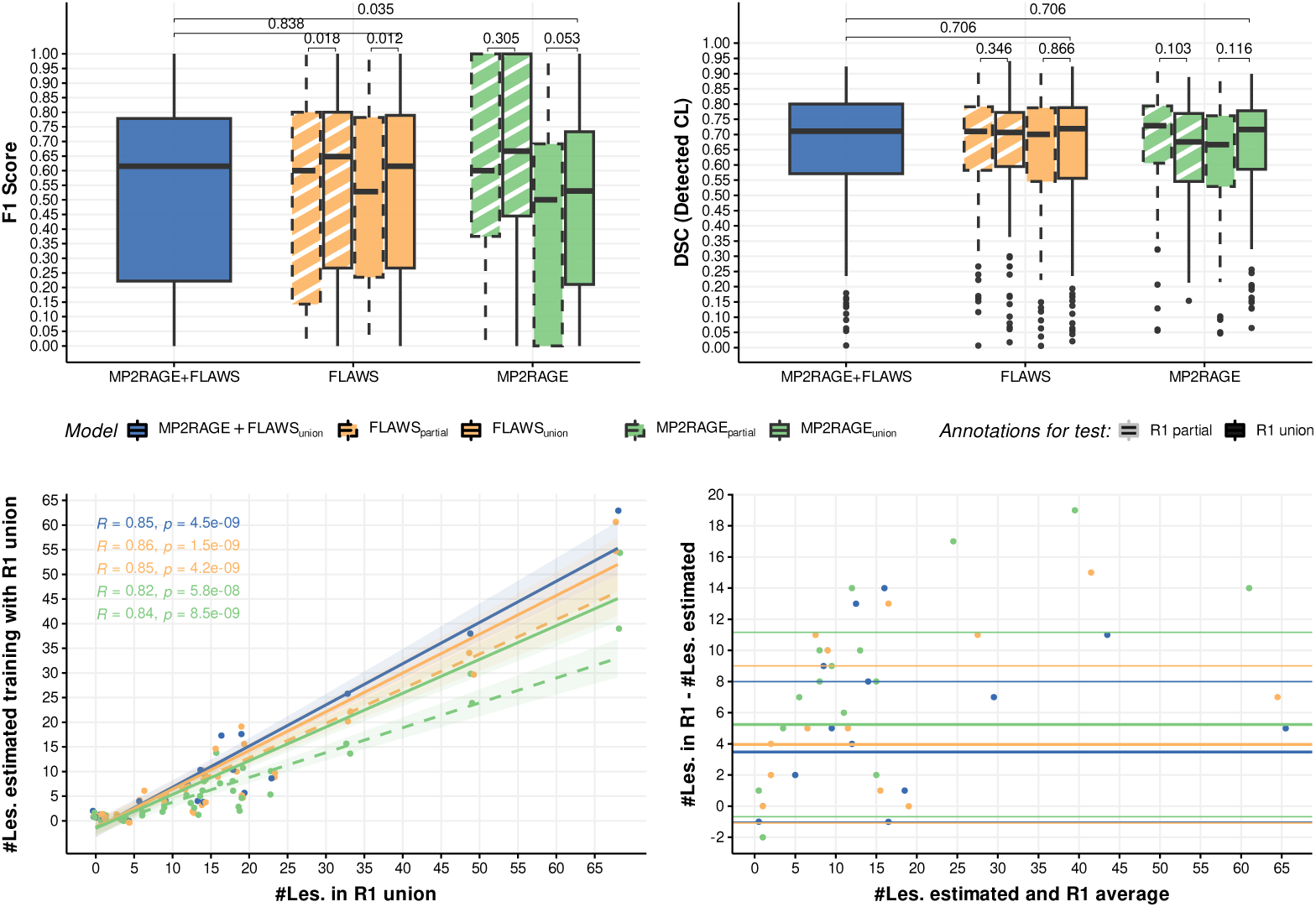
Performance comparison of models trained with different contrast combinations. (Top-Left) Box plots depicting CL detection performance (*F*_1_ score). (Top-Right) Dice Similarity Coefficient (DSC) for overlap assessment of detected CL. The adjusted p-values corresponding with Wilcoxon signed-rank test are shown. (Bottom-Left) Spearman correlation between estimated lesion count and reference standard when training on union annotations and testing on union and partial annotations. *R* coefficients and corresponding p-values are shown for each regression line. (Bottom-Right) Bland-Altman plot showing the agreement between estimated and reference lesion counts for models trained and tested on union annotations. Solid lines indicate models trained and tested on union annotations; dashed lines represent partial annotations for training and stripped patterns for testing.

**Figure 3:**
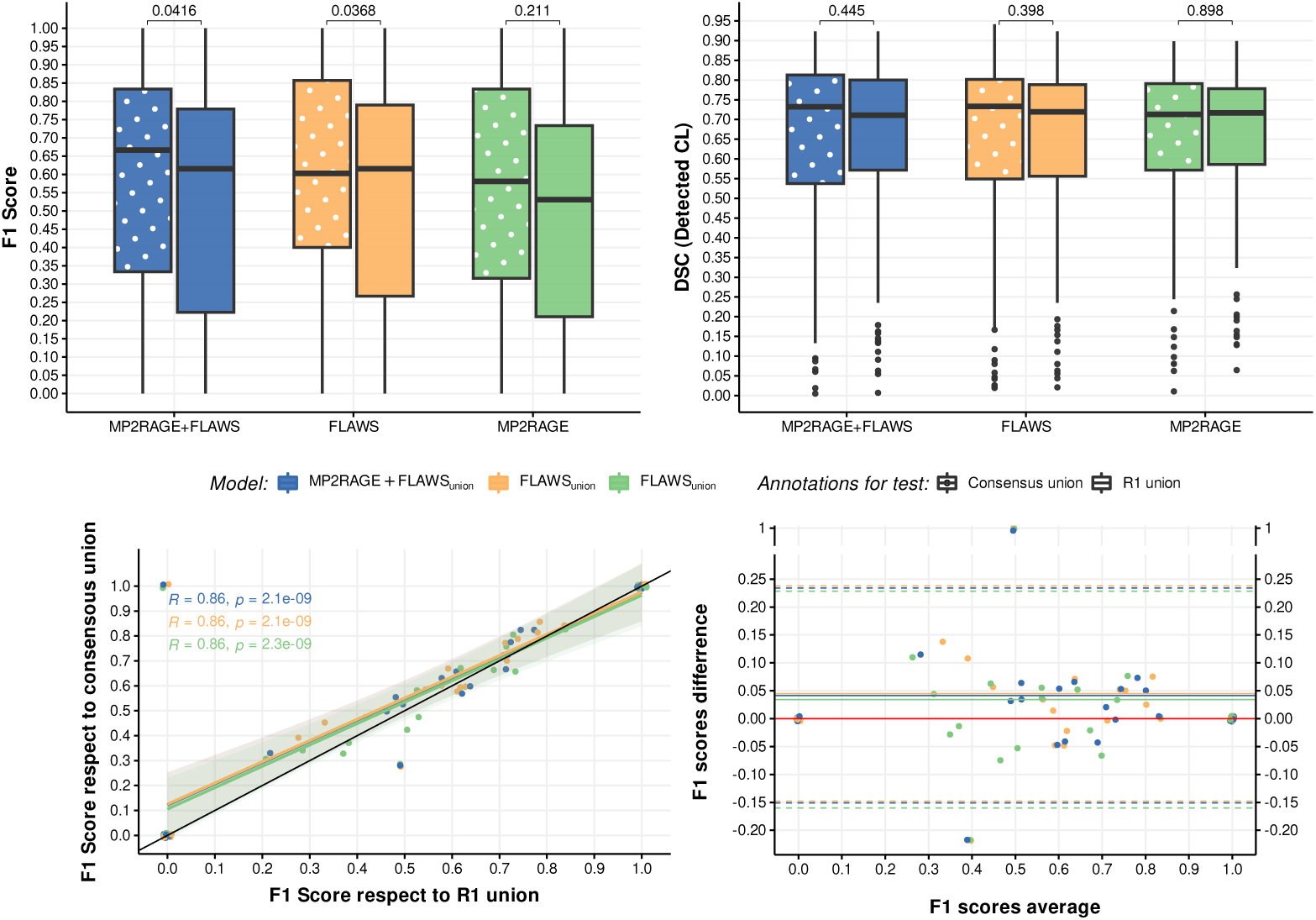
Model performance comparison using single expert (*R1 union*) vs. expert consensus (*consensus union*) annotations. (Top-Left) Box plots depicting CL detection performance (*F*_1_ score). (Top-Right) DSC for overlap assessment of detected CL. (Bottom-Left) Correlation between *F*_1_ scores obtained using *R1 union* and *consensus union* annotations. (Bottom-Right) Bland-Altman plot showing the agreement between *F*_1_ scores from *R1 union* and *consensus union* annotations. Solid box plots represent *R1 union* annotations; dotted box plots represent *consensus union* annotations. *R* coefficients and corresponding p-values are shown for each correlation

DSC analysis (Figure 2 top-right) indicated good overlap between true detected CL across all models (i.e., *MP* 2*RAGE* + *FLAWS_union_* median *DSC* = 0.711 *±* 0.20), with no statistically significant differences observed.

For clarity and to focus on the best-performing models, the correlation and Bland-Altman analyses were conducted only on models trained with *R1 union* annotations. The correlation between detected lesions and the reference standard is illustrated in Figure 2 bottom-left. The MP2RAGE+FLAWS model demonstrated the strongest agreement (*R* = 0.85, *p* = 4.5 *×* 10*^−^*^9^) and similarity with the reference, closely followed by the *FLAWS_union_*model (*R* = 0.86, *p* = 1.5 *×* 10*^−^*^7^). The MP2RAGE-based model showed a lower correlation (*R* = 0.82, *p* = 5.8 *×* 10*^−^*^9^) and a tendency to overestimate CL count.

The Bland-Altman plot (Figure 2 bottom-right) provides further insight into the agreement between the estimated number of lesions and the reference standard. The *MP* 2*RAGE* + *FLAWS_union_* model demonstrates the least bias, with a mean difference close to zero (i.e., 3.48 *±* 4.51 vs 3.97 *±* 5.05 for the *FLAWS* and 5.24 *±* 5.92 for *MP2RAGE*). All models show a tendency to underestimate lesion count, especially in cases with higher lesion loads. The limits of agreement are narrowest for the *MP* 2*RAGE* + *FLAWS* model, indicating better overall agreement with the reference standard across the range of lesion counts.

### 3.2. Experiment 2

This experiment compared model performance when trained and tested on single annotator segmentation maps (*R1 union*) versus testing on expert consensus annotations (*consensus union*).

Analysis of *F*_1_ scores (Figure 3 top-left) revealed improved CL detection for all three input contrast combinations when tested against the *consensus union* compared to the *R1 union*. The *MP* 2*RAGE* + *FLAWS_union_* model showed the most significant improvement, with the median *F*_1_-score increasing from 0.615*±*0.361 to 0.667*±*0.366 (*p* = 0.0416). The *FLAWS_union_* model also demonstrated improvement (*p* = 0.0368), with median *F*_1_ scores increasing from 0.541 *±* 0.36 to 0.585 *±* 0.36 for *R1 union* and *consensus union* annotations, respectively. The *MP* 2*RAGE_union_* model showed a smaller, nonsignificant improvement from mean *F*_1_ = 0.503 *±* 0.36 to *F*_1_ = 0.537 *±* 0.36 (*p* = 0.211).

DSC analysis (Figure 3 top-right) showed no significant differences in overlapping performance for detected lesions between *R1 union* and *consensus union* annotations, suggesting that while the models detect more lesions when compared to the consensus, the accuracy of segmentation remains consistent.

The correlation analysis between *F*_1_ scores obtained using *R1 union* and *consensus union* annotations (Figure 3 bottom-left) demonstrated strong agreement across all models. The *MP* 2*RAGE*+*FLAWS_union_* and *FLAWS_union_*models both showed identical correlation coefficients (*R* = 0.86, *p* = 2.1 *×* 10*^−^*^9^), while the *MP* 2*RAGE_union_* model showed a slightly lower but still strong correlation (*R* = 0.86, *p* = 2.3 *×* 10*^−^*^9^). This high correlation indicates consistent relative performance across annotation methods, regardless of the absolute *F*_1_ scores.

The Bland-Altman plot (Figure 3 bottom-right) provides insight into the systematic differences between *F*_1_ scores obtained using *R1 union* and *consensus union* annotations. All models show a positive mean difference, indicating consistently higher *F*_1_ scores when evaluated against the *consensus union*. The *FLAWS* model demonstrates the largest mean difference (0.045), followed by the *MP* 2*RAGE* + *FLAWS_union_*model (0.041), and the *MP* 2*RAGE_union_* model (0.034). The limits of agreement are equally narrow for the *MP* 2*RAGE* + *FLAWS_union_* and *FLAWS_union_*.

### 3.3. Experiment 3

This experiment evaluated the performance of two models, *MP* 2*RAGE_union_* and *MP* 2*RAGE_partial_* when applied to different test datasets comprising MP2RAGE and MPRAGE sequences. Results are presented in Figure 4.

**Figure 4:**
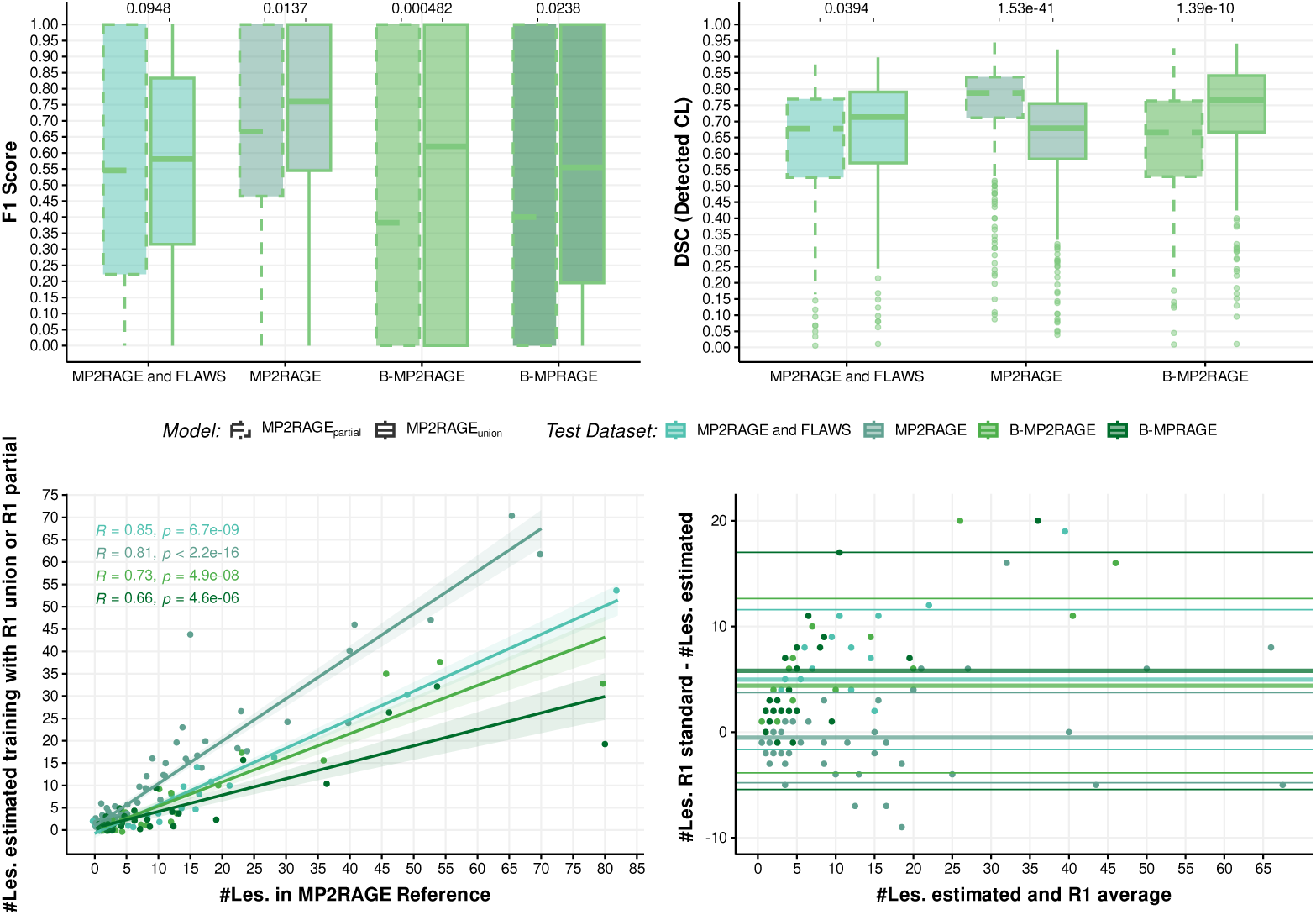
Performance comparison of models trained on MP2RAGE sequence employing annotations derived from MP2RAGE alone (*R1 partial*) or all contrasts (*R1 union*), evaluated on external research (*B-MP2RAGE*), clinical (*B-MPRAGE*), and on MP2RAGE images of internal datasets (*A-MP2RAGE and FLAWS* and *A-MP2RAGE*). (Top-Left) Box plots depicting CL detection performance (*F*_1_ score). (Top-Right) Dice Similarity Co-efficient (DSC) for overlap assessment of detected CL. Adjusted p-values from Wilcoxon signed-rank tests are shown. (Bottom-Left) Spearman correlation between estimated lesion count and reference standard for different datasets and training annotations. (Bottom-Right) Bland-Altman plot showing the agreement between estimated and reference lesion counts for the *MP* 2*RAGE_union_* model trained with R1 union annotations. Solid lines indicate models trained with R1 union annotations; dashed lines represent R1 partial annotations for training.

**Figure 5:**
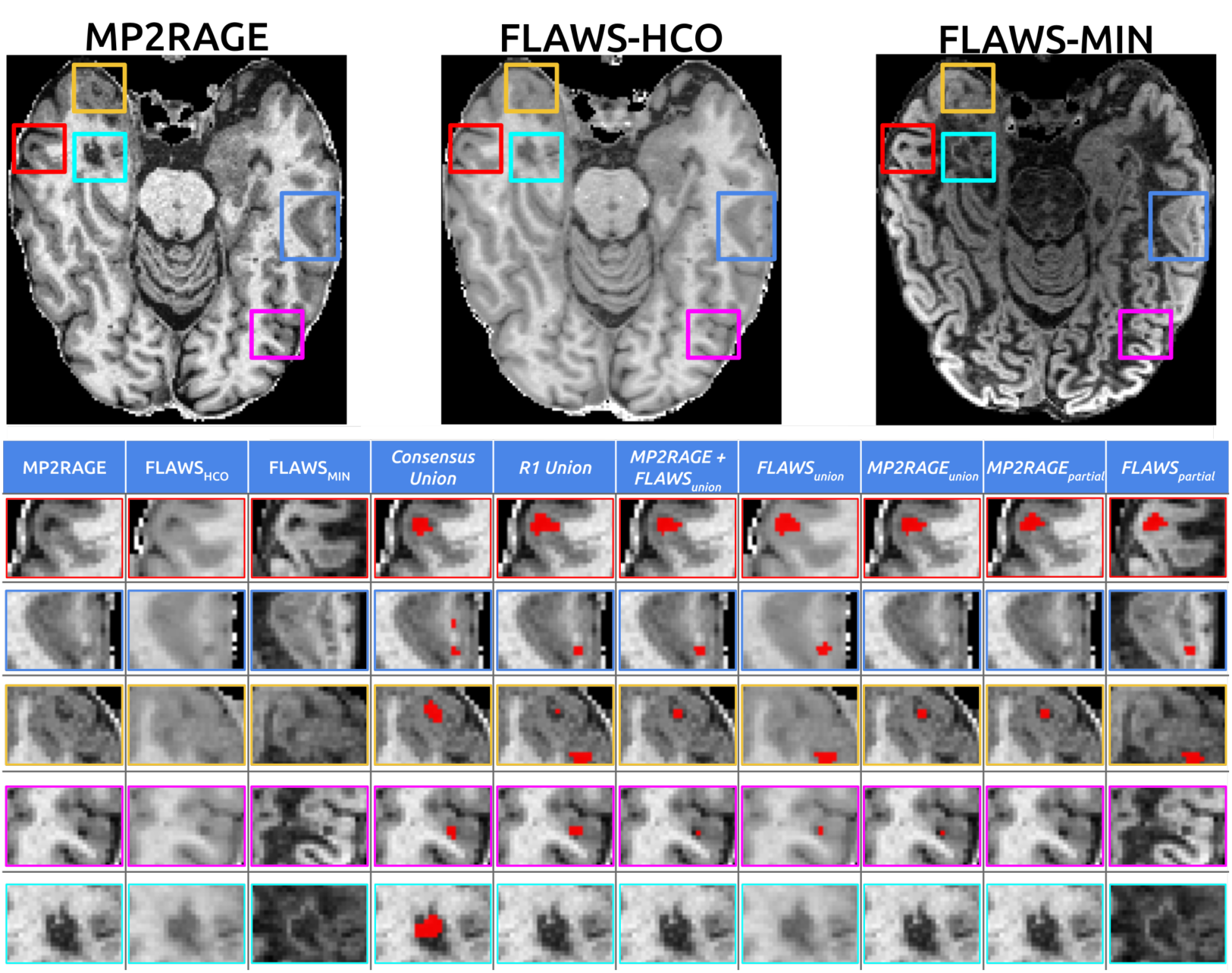
Examples of different CL across available contrasts (top and zoomed views in the first three grid columns), manual segmentations (*Consensus union* and *R1 Union* columns), and predictions from each model (Model Development) in the last five grid columns.

Analysis of *F*_1_ scores (Figure 4 top-left) revealed that the *MP* 2*RAGE_union_* model, trained using annotations derived from all available sequences, consistently outperformed the *MP* 2*RAGE_partial_* model in lesion detection across all test datasets. This performance advantage was particularly pronounced in the external dataset, *B-MP2RAGE*, where *MP* 2*RAGE_union_*achieved a median *F*_1_ score of 0.621*±*0.406 compared to 0.383*±*0.419 for *MP* 2*RAGE_partial_* (*p* = 4.82 *×* 10*^−^*^4^).

Importantly, for the clinical MPRAGE sequence (*B-MPRAGE* external testing dataset), the *MP* 2*RAGE_union_* model obtained a median *F*_1_ score of 0.55 *±* 0.356, while its partial counterpart achieved only 0.40 *±* 0.44).

Notably, the *MP* 2*RAGE_union_* model’s performance on *B-MP2RAGE* was comparable to its performance on the held-out subjects from the internal *A-MP2RAGE and FLAWS* dataset (*F*_1_ = 0.581 *±* 0.36), suggesting robust generalization to external data.

The *MP* 2*RAGE_union_*model demonstrated superior detection capabilities on the internal test dataset *MP2RAGE*, achieving a median *F*_1_ score of 0.724 *±* 0.34.

DSC analysis (Figure 4 top-right) revealed a more nuanced picture of segmentation accuracy. Interestingly, the *MP* 2*RAGE_partial_* model showed better DSC results for the internal *MP2RAGE* dataset, a trend not observed in the other test datasets.

The correlation analysis (Figure 4 bottom-left) provides insight into the relationship between estimated and reference lesion counts across datasets. The *MP* 2*RAGE_union_* model showed strong correlations for both the internal *A-MP2RAGE* dataset (*R* = 0.81, *p <* 2.2 *×* 10*^−^*^16^) and the *A-MP2RAGE and FLAWS* (*R* = 0.85, *p* = 567 *×* 10*^−^*^09^). The correlation was weaker for the external *B-MP2RAGE* dataset (*R* = 0.73, *p* = 4.9 *×* 10*^−^*^08^) and *B-MPRAGE* (*R* = 0.66, *p* = 4.6 *×* 10*^−^*^06^),

The Bland-Altman plot (Figure 4 bottom-right), focusing solely on the *MP* 2*RAGE_union_*model trained with *R1 union* annotations, demonstrates the agreement between estimated and reference lesion counts.

### 3.4. Qualitative results

Figure 5 presents segmentation predictions for each model (last five columns in the grid) across various lesion types. The figure illustrates several key observations: In the first row (red square), all models and manual segmentations concur on lesion identification. The second row (dark blue rectangle) showcases a case where the *Consensus Union*annotations reveal a previously unidentified lesion. Models relying solely on MP2RAGE input fail to detect both lesions highlighted in the revision, while FLAWS-based models identify one lesion, consistent with the *R1 Union* annotation, regardless of the training annotation type. Conversely, the third row (yellow square) demonstrates a scenario where MP2RAGE-based models align with the consensus manual annotation, detecting one lesion, while FLAWS-based models identify two lesions as per the *R1 Union* reference. The fourth row (violet square) highlights the performance of models utilizing all available annotation information. Only models trained on *R1 union* annotations correctly identify the small lesion presented, in concordance with Experiment 2. The final row illustrates a case where a lesion, initially undetected in the single manual annotation, was subsequently missed by all models.

Notably, the *MP* 2*RAGE* + *FLAWS_union_* model consistently shows the closest alignment with the consensus annotations across various scenarios, corroborating the quantitative findings from previous experiments.

## 4. Discussion

This study demonstrates the potential of leveraging FLAWS-derived contrasts for improved automatic CL detection and segmentation in MS patients using DL models. Our findings have several important implications for the field of MS MRI imaging and AI-assisted diagnosis.

### Enhanced CL DL Detection with FLAWS

The integration of FLAWS-derived contrasts significantly improved automatic CL assessment, as demonstrated in Experiment 1 and Experiment 2. Models incorporating FLAWS (*MP* 2*RAGE*+*FLAWS_union_* and *FLAWS_union_*) consistently outperformed the *MP* 2*RAGE_union_* model, with higher *F*_1_ scores and stronger correlations to reference standards. This performance enhancement underscores the value of FLAWS in CL detection and segmentation.

Furthermore, comparative analysis of *FLAWS_partial_*-*FLAWS_union_* and *MP* 2*RAGE_partial_*-*MP* 2*RAGE_union_* pairs against their respective *R1 partial* or *R1 union* test sets revealed that models trained on union annotations derived from all available contrasts consistently outperformed those trained on partial annotations. This finding suggests that the comprehensive annotation strategy enhances model performance without introducing contrast-specific biases.

These results collectively address our first objective, demonstrating that FLAWS enriches automatic CL assessment through both its novel contrast properties and the derived annotations.

### Enhanced Sensitivity of Models in CL Detection

Experiment 2 addressed our second objective by evaluating the models’ ability to detect CL potentially overlooked by individual raters. This is particularly relevant given the well-known challenges in CL identification, which depend on both sequence sensitivity and rater expertise [7].

We compared model performance using single-rater annotations against consensus-reviewed annotations. Notably, models, even when trained on single-rater annotations (*MP* 2*RAGE* + *FLAWS_union_*and *FLAWS_union_*), demonstrated statistically significant better *F*_1_ scores when evaluated on consensus annotations than on the single-rater annotations in the test set. This suggests that our models can potentially identify CL missed by individual expert raters while maintaining high segmentation accuracy (as indicated by consistent DSC values), showing the model’s ability to extract relevant information beyond the standard exactness available at training. These findings are particularly promising considering the limited dataset size, a common challenge when introducing new imaging sequences. The models’ ability to generalize and potentially outperform single-rater annotations highlights the robustness of the DL approach in CL detection.

### Generalization and Transferability to Clinical sequences

Experiment 3 addressed our third objective by evaluating the generalization capabilities of models trained on FLAWS-derived annotations when applied to MP2RAGE and MPRAGE, including both internal and external testing datasets. The *MP* 2*RAGE_union_* model, trained on annotations derived from all available contrasts, demonstrated superior performance across different datasets.

Notably, the *MP* 2*RAGE_union_* model achieved higher *F*_1_ scores compared to *MP* 2*RAGE_partial_*on both internal and external MP2RAGE sequence based datasets. This performance advantage was particularly pronounced on the external, shifted datasets: the *B-MP2RAGE* dataset (median *F*_1_ = 0.621 *±* 0.406 vs. 0.383 *±* 0.419, *p* = 4.82 *×* 10*^−^*^4^, *p* = 4.82 *×* 10*^−^*^4^), and the *B-MPRAGE* dataset (median *F*_1_ = 0.55 *±* 0.356 vs. 0.40 *±* 0.44, *p* = 0.02). These results suggest a robust model capability for generalization, even under significantly shifted data, as in the case of the clinical MPRAGE sequence.

Interestingly, the model’s performance on the internal *MP2RAGE* dataset (94 previously unseen subjects) surpassed its performance on the held-out subjects for internal testing from the *MP2RAGE and FLAWS* dataset. While this finding may seem counterintuitive initially, it can be attributed to the fact that the reference annotations for these datasets were derived solely from the MP2RAGE contrast. This observation reinforces the model’s ability to effectively leverage information learned from FLAWS-derived annotations, even when applied to single-contrast data.

Furthermore, the model’s ability to maintain or even improve performance on MP2RAGE-only data underscores the value of incorporating FLAWS-derived contrasts in the training process. Our results indicate that advanced imaging techniques like FLAWS can enhance CL detection in settings where such sequences are not available, thereby potentially bridging the gap between research capabilities and clinical practice in CL detection through label transfer knowledge.

### Limitations and Future Directions

While our study yields promising results, it is crucial to acknowledge its limitations. The training set, though realistically sized for a novel imaging biomarker study, is relatively small. This, coupled with the lack of publicly available CL datasets—a consequence of CL recent emergence as an MS imaging biomarker—presents challenges for comprehensive model development and validation.

Our qualitative results also suggest potential inconsistencies in CL annotation criteria between individual raters and consensus reviews (as seen in the last row of Qualitative results). This variability in lesion definition, akin to what machine learning literature terms *concept shift*, highlights the need for standardized annotation protocols, especially for multi-center studies [29].

To address these limitations and further advance the field, we propose several directions for future research. Validation on larger multi-center datasets will be crucial to rigorously assess model generalizability across diverse patient populations and imaging protocols. The development of standardized, publicly available CL datasets would facilitate benchmarking and collaborative research in this emerging field. To facilitate this transition and promote further research, we have made our trained models available through DockerHub, aiming to accelerate the development and validation of AI-assisted CL detection tools across different clinical and research environments.

### Clinical Impact

This work addresses a critical need in MS diagnosis and monitoring, as CL are increasingly recognized as important indicators of disease progression [5, 6]. The models’ demonstrated ability to align with consensus annotations while maintaining performance across different sequences suggests potential for practical clinical implementation. Furthermore, our findings substantially contribute to the growing body of evidence supporting the adoption of AI-assisted tools in radiology, particularly for complex tasks such as CL assessment where expert consensus may be impractical or time-consuming.

### Conclusion

This study demonstrates the significant potential of integrating advanced MRI sequences, specifically FLAWS, with state-of-the-art DL models to enhance CL detection and segmentation in MS. Our work not only advances the technical capabilities of CL detection but also paves the way for more accurate, consistent, and efficient MS assessment through the synergistic combination of novel imaging sequences and AI-assisted analysis.

## Data Availability

The authors do not have permission to share the data

## References

[1] M. Filippi, P. Preziosa, B. L. Banwell, F. Barkhof, O. Ciccarelli, N. De Stefano, J. J. G. Geurts, F. Paul, D. S. Reich, A. T. Toosy, A. Traboulsee, M. P. Wattjes, T. A. Yousry, A. Gass, C. Lubetzki, B. G. Weinshenker, M. A. Rocca, Assessment of lesions on magnetic resonance imaging in multiple sclerosis: practical guidelines, Brain 142 (7) (2019) 1858–1875. doi:10.1093/brain/awz144. URL 10.1093/brain/awz144

[2] C. Granziera, J. Wuerfel, F. Barkhof, M. Calabrese, N. De Stefano, C. Enzinger, N. Evangelou, M. Filippi, J. J. G. Geurts, D. S. Reich, M. A. Rocca, S. Ropele, A. Rovira, P. Sati, A. T. Toosy, H. Vrenken, C. A. M. Gandini Wheeler-Kingshott, L. Kappos, the MAGNIMS Study Group, Quantitative magnetic resonance imaging towards clinical application in multiple sclerosis, Brain 144 (5) (2021) 1296–1311. doi:10.1093/brain/awab029. URL 10.1093/brain/awab029

[3] A. J. Thompson, B. L. Banwell, F. Barkhof, W. M. Carroll, T. Coetzee, G. Comi, J. Correale, F. Fazekas, M. Filippi, M. S. Freedman, K. Fujihara, S. L. Galetta, H. P. Hartung, L. Kappos, F. D. Lublin, R. A. Marrie, A. E. Miller, D. H. Miller, X. Montalban, E. M. Mowry, P. S. Sorensen, M. Tintoré, A. L. Traboulsee, M. Trojano, B. M. Uitdehaag, S. Vukusic, E. Waubant, B. G. Weinshenker, S. C. Reingold, J. A. Cohen, Diagnosis of multiple sclerosis: 2017 revisions of the McDonald criteria, The Lancet Neurology 17 (2) (2018) 162–173, publisher: Elsevier. doi:10.1016/S1474-4422(17)30470-2.

[4] M. Filippi, M. A. Rocca, M. A. Horsfield, S. Hametner, J. J. G. Geurts, G. Comi, H. Lassmann, Imaging Cortical Damage and Dysfunction in Multiple Sclerosis, JAMA Neurology 70 (5) (2013) 556–564. doi:10.1001/jamaneurol.2013.1954. URL 10.1001/jamaneurol.2013.1954

[5] E. S. Beck, W. A. Mullins, J. dos Santos Silva, S. Filippini, P. Parvathaneni, J. Maranzano, M. Morrison, D. J. Suto, C. Donnay, H. Dieckhaus, N. J. Luciano, K. Sharma, M. I. Gaitán, J. Liu, J. A. de Zwart, P. van Gelderen, I. Cortese, S. Narayanan, J. H. Duyn, G. Nair, P. Sati, D. S. Reich, Contribution of new and chronic cortical lesions to disability accrual in multiple sclerosis, Brain Communications (2024) fcae158doi:10.1093/braincomms/fcae158. URL 10.1093/braincomms/fcae158

[6] A. Cagol, R. Cortese, M. Barakovic, S. Schaedelin, E. Ruberte, M. Absinta, F. Barkhof, M. Calabrese, M. Castellaro, O. Ciccarelli, S. Cocozza, N. De Stefano, C. Enzinger, M. Filippi, M. Jurynczyk, P. Maggi, N. Mahmoudi, S. Messina, X. Montalban, J. Palace, G. Pontillo, A.-K. Pröbstel, M. A. Rocca, S. Ropele, A. Rovira, M. M. Schoonheim, P. Sowa, E. Strijbis, M. P. Wattjes, M. P. Sormani, L. Kappos, C. Granziera, MAGNIMS Study Group, Diagnostic Performance of Cortical Lesions and the Central Vein Sign in Multiple Sclerosis, JAMA Neurology 81 (2) (2024) 143–153. doi:10.1001/jamaneurol.2023.4737. URL 10.1001/jamaneurol.2023.4737

[7] M. A. J. Madsen, V. Wiggermann, S. Bramow, J. R. Christensen, F. Sellebjerg, H. R. Siebner, Imaging cortical multiple sclerosis lesions with ultra-high field MRI, NeuroImage: Clinical 32 (2021) 102847. doi:10.1016/j.nicl.2021.102847. URL https://www.sciencedirect.com/science/article/pii/ S2213158221002916

[8] J. Maranzano, M. Dadar, D. Rudko, D. De Nigris, C. Elliott, J. Gati, S. Morrow, R. Menon, D. Collins, D. Arnold, S. Narayanan, Comparison of Multiple Sclerosis Cortical Lesion Types Detected by Multicontrast 3T and 7T MRI, AJNR: American Journal of Neuroradiology 40 (7) (2019) 1162–1169. doi:10.3174/ajnr.A6099. URL https://www.ncbi.nlm.nih.gov/pmc/articles/PMC7048547/

[9] G. Hinton, Deep Learning—A Technology With the Potential to Transform Health Care, JAMA 320 (11) (2018) 1101. doi:10.1001/jama.2018.11100.

[10] J. Müller, F. La Rosa, J. Beaumont, C. Tsagkas, R. Rahmanzadeh, M. Weigel, M. Bach Cuadra, G. Gambarota, C. Granziera, Fluid and White Matter Suppression : New Sensitive 3 T Magnetic Resonance Imaging Contrasts for Cortical Lesion Detection in Multiple Sclerosis, Investigative Radiology 57 (9) (2022) 592–600. doi:10.1097/RLI.0000000000000877.

[11] A. S. Nielsen, R. P. Kinkel, E. Tinelli, T. Benner, J. Cohen-Adad, C. Mainero, Focal cortical lesion detection in multiple sclerosis: 3 tesla DIR versus 7 tesla FLASH-T2*, Journal of Magnetic Resonance Imaging 35 (3) (2012) 537–542, _eprint: https://onlinelibrary.wiley.com/doi/pdf/10.1002/jmri.22847. doi:10.1002/jmri.22847. URL https://onlinelibrary.wiley.com/doi/abs/10.1002/jmri. 22847

[12] M. Tanner, G. Gambarota, T. Kober, G. Krueger, D. Erritzoe, J. P. Marques, R. Newbould, Fluid and white matter suppression with the MP2RAGE sequence, Journal of magnetic resonance imaging: JMRI 35 (5) (2012) 1063–1070. doi:10.1002/jmri.23532.

[13] F. La Rosa, A. Abdulkadir, M. J. Fartaria, R. Rahmanzadeh, P. J. Lu, R. Galbusera, M. Barakovic, J. P. Thiran, C. Granziera, M. B. Cuadra, Multiple sclerosis cortical and WM lesion segmentation at 3T MRI: a deep learning method based on FLAIR and MP2RAGE, NeuroImage: Clinical 27 (2020) 102335, publisher: Elsevier. doi:10.1016/J.NICL. 2020.102335.

[14] F. La Rosa, E. S. Beck, J. Maranzano, R.-A. Todea, P. van Gelderen, J. A. de Zwart, N. J. Luciano, J. H. Duyn, J.-P. Thiran, C. Granziera, D. S. Reich, P. Sati, M. Bach Cuadra, Multiple sclerosis cortical lesion detection with deep learning at ultra-high-field MRI, NMR in Biomedicine 35 (8) (2022) e4730. doi:10.1002/nbm.4730. URL https://onlinelibrary.wiley.com/doi/abs/10.1002/nbm. 4730

[15] F. Spagnolo, A. Depeursinge, S. Schädelin, A. Akbulut, H. Müller, M. Barakovic, L. Melie-Garcia, M. Bach Cuadra, C. Granziera, How far MS lesion detection and segmentation are integrated into the clinical workflow? A systematic review, NeuroImage: Clinical 39 (2023) 103491. doi:10.1016/j.nicl.2023.103491. URL https://www.sciencedirect.com/science/article/pii/ S2213158223001821

[16] F. Isensee, P. F. Jaeger, S. A. Kohl, J. Petersen, K. H. Maier-Hein, nnU-Net: a self-configuring method for deep learning-based biomedical image segmentation, Nature Methods 18 (2) (2021) 203–211, publisher: Nature Research. doi:10.1038/s41592-020-01008-z.

[17] B. B. Avants, N. Tustison, H. Johnson, Advanced Normalization Tools (ANTS), Insight Journal 2 (365) (2009) 1–35.

[18] A. Hoopes, J. S. Mora, A. V. Dalca, B. Fischl, M. Hoffmann, SynthStrip: skull-stripping for any brain image, NeuroImage 260 (2022) 119474. doi: 10.1016/j.neuroimage.2022.119474. URL https://www.ncbi.nlm.nih.gov/pmc/articles/PMC9465771/

[19] P. A. Yushkevich, J. Piven, H. C. Hazlett, R. G. Smith, S. Ho, J. C. Gee, G. Gerig, User-guided 3D active contour segmentation of anatomical structures: significantly improved efficiency and reliability, NeuroImage 31 (3) (2006) 1116–1128. doi:10.1016/j.neuroimage.2006.01.015.

[20] M. J. Fartaria, G. Bonnier, A. Roche, T. Kober, R. Meuli, D. Rotzinger, R. Frackowiak, M. Schluep, R. Du Pasquier, J.-P. Thiran, G. Krueger, M. Bach Cuadra, C. Granziera, Automated detection of white matter and cortical lesions in early stages of multiple sclerosis, Journal of Magnetic Resonance Imaging 43 (6) (2016) 1445–1454, _eprint: https://onlinelibrary.wiley.com/doi/pdf/10.1002/jmri.25095. doi:10.1002/jmri.25095. URL https://onlinelibrary.wiley.com/doi/abs/10.1002/jmri.25095

[21] D. P. Kingma, J. Ba, Adam: A Method for Stochastic Optimization, arXiv: 1412.6980 (Dec. 2014). URL http://arxiv.org/abs/1412.6980

[22[ F. Kofler, S. Shit, I. Ezhov, L. Fidon, I. Horvath, R. Al-Maskari, H. Li, H. Bhatia, T. Loehr, M. Piraud, A. Erturk, J. Kirschke, J. C. Peeken, T. Vercauteren, C. Zimmer, B. Wiestler, B. Menze, blob loss: instance imbalance aware loss functions for semantic segmentation, arXiv:2205.08209 [cs, eess] (Jun. 2023). doi:10.48550/arXiv.2205.08209. URL http://arxiv.org/abs/2205.08209

[23] C. H. Sudre, K. Van Wijnen, F. Dubost, H. Adams, D. Atkinson, F. Barkhof, M. A. Birhanu, E. E. Bron, R. Camarasa, N. Chaturvedi, Y. Chen, Z. Chen, S. Chen, Q. Dou, T. Evans, I. Ezhov, H. Gao, M. Girones Sanguesa, J. D. Gispert, B. Gomez Anson, A. D. Hughes, M. A. Ikram, S. Ingala, H. R. Jaeger, F. Kofler, H. J. Kuijf, D. Kutnar, M. Lee, B. Li, L. Lorenzini, B. Menze, J. L. Molinuevo, Y. Pan, E. Puybareau, R. Rehwald, R. Su, P. Shi, L. Smith, T. Tillin, G. Tochon, H. Urien, B. H. M. van der Velden, I. F. van der Velpen, B. Wiestler, F. J. Wolters, P. Yilmaz, M. de Groot, M. W. Vernooij, M. de Bruijne, Where is VALDO? VAscular Lesions Detection and segmentatiOn challenge at MICCAI 2021, Medical Image Analysis 91 (2024) 103029. doi:10.1016/j.media.2023.103029. URL https://www.sciencedirect.com/science/article/pii/ S136184152300289X

[24] D. C. Castro, I. Walker, B. Glocker, Causality matters in medical imaging, Nature Communications 11 (1) (2020) 1–10, publisher: Nature Research. doi:10.1038/s41467-020-17478-w

[25] L. Maier-Hein, A. Reinke, P. Godau, M. D. Tizabi, F. Buettner, E. Christodoulou, B. Glocker, F. Isensee, J. Kleesiek, M. Kozubek, M. Reyes, M. A. Riegler, M. Wiesenfarth, A. E. Kavur, C. H. Sudre, M. Baumgartner, M. Eisenmann, D. Heckmann-Nötzel, T. Rädsch, L. Acion, M. Antonelli, T. Arbel, S. Bakas, A. Benis, M. B. Blaschko, M. J. Cardoso, V. Cheplygina, B. A. Cimini, G. S. Collins, K. Farahani, L. Ferrer, A. Galdran, B. van Ginneken, R. Haase, D. A. Hashimoto, M. M. Hoffman, M. Huisman, P. Jannin, C. E. Kahn, D. Kainmueller, B. Kainz, A. Karargyris, A. Karthikesalingam, F. Kofler, A. Kopp-Schneider, A. Kreshuk, T. Kurc, B. A. Landman, G. Litjens, A. Madani, K. Maier-Hein, A. L. Martel, P. Mattson, E. Meijering, B. Menze, K. G. M. Moons, H. Müller, B. Nichyporuk, F. Nickel, J. Petersen, N. Rajpoot, N. Rieke, J. Saez-Rodriguez, C. I. Sánchez, S. Shetty, M. van Smeden, R. M. Summers, A. A. Taha, A. Tiulpin, S. A. Tsaftaris, B. Van Calster, G. Varoquaux, P. F. Jäger, Metrics reloaded: recommendations for image analysis validation, Nature Methods 21 (2) (2024) 195–212, publisher: Nature Publishing Group. doi:10.1038/s41592-023-02151-z. URL https://www.nature.com/articles/s41592-023-02151-z

[26] G. Lehmann, Label object representation and manipulation with ITK, Tech. rep., Kitware, publication Title: The Insight Journal Volume: 8 (2007). URL http://hdl.handle.net/1926/584

27. R. C. Team, R: A Language and Environment for Statistical Computing (2021). URL https://www.R-project.org/

28. A. Kassambara, rstatix: Pipe-Friendly Framework for Basic Statistical Tests (2023). URL https://CRAN.R-project.org/package=rstatix

29. B. Nichyporuk, J. Cardinell, J. Szeto, R. Mehta, J.-P. R. Falet, D. L. Arnold, S. A. Tsaftaris, T. Arbel, Rethinking Generalization: The Impact of Annotation Style on Medical Image Segmentation, arXiv:2210.17398 [cs, eess] (Dec. 2022). doi:10.48550/arXiv.2210.17398. URL http://arxiv.org/abs/2210.17398

